# Pregnancy-Associated Bleeding and Genetics: Five Sequence Variants in the Myometrium and Progesterone Signaling Pathway are associated with postpartum hemorrhage

**DOI:** 10.1101/2023.08.10.23293932

**Authors:** David Westergaard, Valgerdur Steinthorsdottir, Lilja Stefansdottir, Palle Duun Rohde, Xiaoping Wu, Frank Geller, Jaakko Tyrmi, Aki S. Havulinna, Pol Sole Navais, Christopher Flatley, Sisse Rye Ostrowski, Ole Birger Pedersen, Christian Erikstrup, Erik Sørensen, Christina Mikkelsen, Mie Topholm Brun, Bitten Aagaard Jensen, Thorsten Brodersen, Henrik Ullum, FinnGen, Danish Blood Donor Study Genomic Consortium, Estonian Biobank Research Team, Nordic Collaboration for Womens and Reproductive Health, Per Magnus, Ole A. Andreassen, Pål R. Njolstad, Astrid Marie Kolte, Lone Krebs, Mette Nyegaard, Thomas Folkmann Hansen, Bjarke Fenstra, Mark Daly, Cecilia M. Lindgren, Gudmar Thorleifsson, Olafur A. Stefansson, Gardar Sveinbjornsson, Daniel F. Gudbjartsson, Unnur Thorsteinsdottir, Karina Banasik, Bo Jacobsson, Triin Laisk, Hannele Laivuori, Kari Stefansson, Søren Brunak, Henriette Svarre Nielsen

## Abstract

Bleeding in early pregnancy and postpartum hemorrhage (PPH) bear substantial risks, with the former closely associated with pregnancy loss and the latter being the foremost cause of maternal death, underscoring the severity of these complications in maternal-fetal health. Here, we investigated the genetic variation underlying aspects of pregnancy-associated bleeding and identified five loci associated with PPH through a meta-analysis of 21,512 cases and 259,500 controls. Functional annotation analysis indicated candidate genes, *HAND2*, *TBX3*, and *RAP2C*/*FRMD7,* at three loci and showed that at each locus, associated variants were located within binding sites for progesterone receptors (PGR). Furthermore, there were strong genetic correlations with birth weight, gestational duration, and uterine fibroids. Early bleeding during pregnancy (28,898 cases and 302,894 controls) yielded no genome-wide association signals, but showed strong genetic correlation with a variety of human traits, indicative of polygenic and pleiotropic effects. Our results suggest that postpartum bleeding is related to myometrium dysregulation, whereas early bleeding is a complex trait related to underlying health and possibly socioeconomic status.

## Introduction

Pregnancy-associated bleeding can occur at all stages of pregnancy. Bleeding in early pregnancy can range in significance from a benign event with no adverse effects, to an indication of ongoing pregnancy loss, and even serve as a potential marker for later pregnancy loss, obstetric complications, and long-term maternal comorbidities^1, 2^. Postpartum hemorrhage (PPH) is the leading cause of maternal mortality, with approximately 100,000 young and otherwise healthy women dying every year worldwide^3^. Despite affecting more than one in ten births and being a heritable condition, PPH remains unexplored at the genetic and molecular level^4^. Prior candidate gene studies have focused on genes involved in the coagulation pathways^5^. Even though the etiology of PPH is multifactorial, it often occurs even when established risk factors are not present^6, 7^.

The primary cause of PPH is uterine atony, which accounts for 70% of all cases^3^. Other causes include retained placental tissue, trauma, and congenital or acquired coagulation disorders. Early identification and correct management of PPH can prevent maternal mortality and morbidity^8^. Therefore, there is great interest in assessing PPH risk prior to labor, and a large body of literature has described detailed prognostic models. However, a recent review showed that almost half of the existing prognostic models include features that can only be obtained postpartum^9^. Consequently, there is an urgent clinical need to understand the molecular etiology and identify novel biomarkers that characterize high-risk women prior to labor to initiate timely preventive measures and monitoring.

Here, we report the results of genome-wide association studies (GWAS) of up to 302,894 women from six Northern European cohorts to identify the genetic etiology of bleeding during different stages of pregnancy. Our results reveal complexity in the genetics of early bleeding and highlight the importance of the myometrium and progesterone-responsive genes in the etiology of PPH.

## Results

### Overall findings

Combining data from six Northern European cohorts including up to 331,792 women we investigated the genetic architecture of three phenotypes related to bleeding during pregnancy; early bleeding (28,898 cases), antepartum hemorrhage (3,236 cases), and postpartum hemorrhage (PPH) (21,521 cases) (Figure 1A). We further divided early bleeding into “early bleeding with any outcome” (28,898 cases) and “early bleeding ending in live birth” (6,356 cases) (Supplementary Table 1). We included up to 18,009,056 sequence variants in a meta-analysis and identified five loci (chromosome 4, 6, 10, 12, and X) that were associated with PPH using a functionally informed multiple testing correction (Figure 2B, Table 1). The effect sizes were similar across all cohorts (Supplementary Figure 1A), and conditional analysis revealed no secondary signals. We observed no significant associations for early bleeding and antepartum hemorrhage (Supplementary Figure 2-4). In addition, we analyzed uterine atony (13,048 cases and 261,809 controls) and retained placental tissue (6,256 cases and 266,427 controls), where three (chromosome 4, 6, and 10) and one (chromosome X) of the five associated loci passed multiple testing correction, respectively (Figure 1C, Supplementary Table 2). We did not observe any significant differences in effect sizes between uterine atony and retained placental tissue, when comparing the lead variants from the five loci (Supplementary Table 2). We found no evidence of confounding or inflation in any of the analyses (Supplementary Table 3).

**Figure 1.**
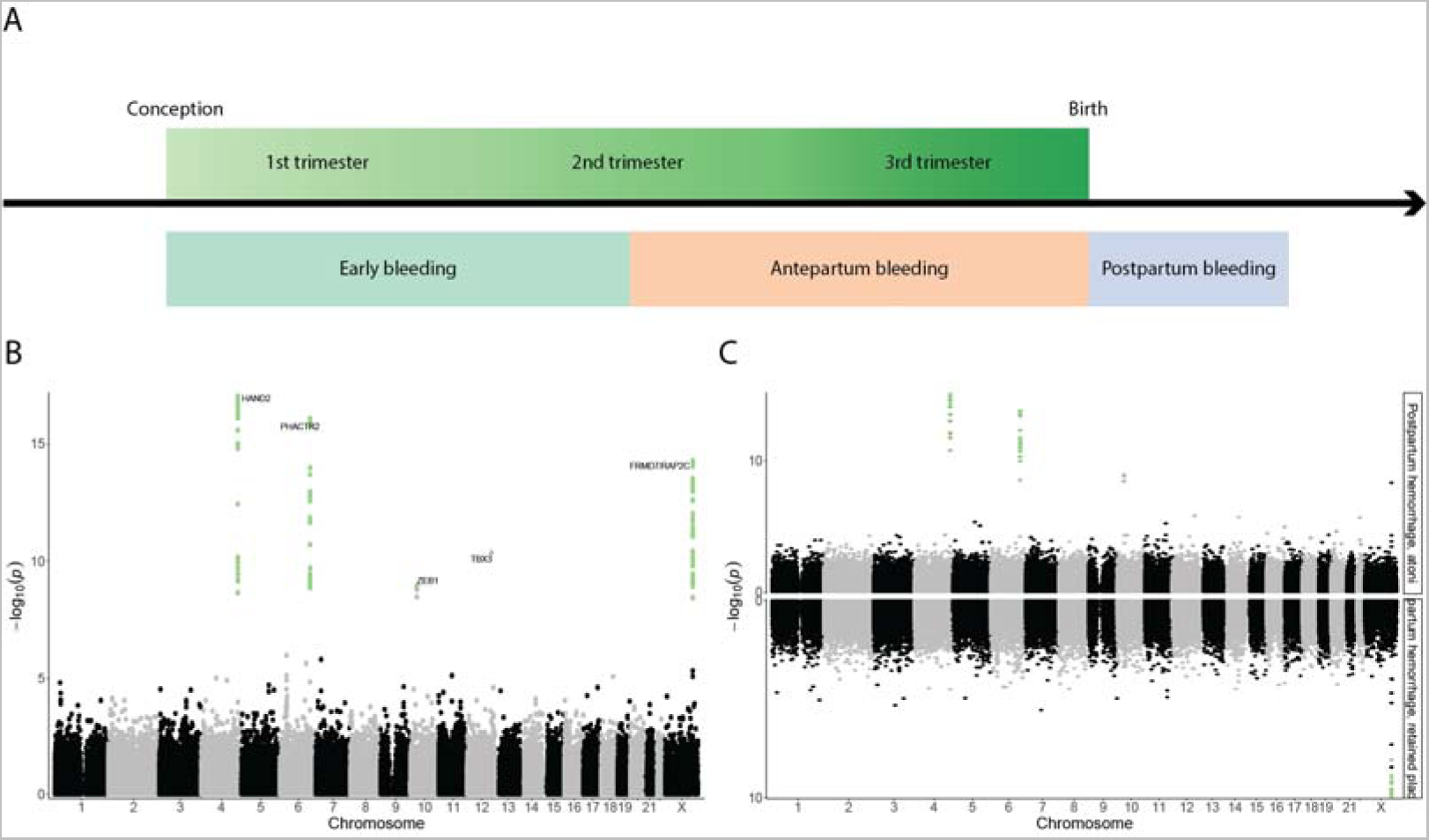
(A) Overview of the phenotypes under investigation. Early bleeding occurs up and until the 20th gestational week, antepartum between the 20th gestational week and birth, and postpartum hemorrhage after birth. (B) Manhattan plot of postpartum hemorrhage showing the 18M variants, with SNPs passing the functionally informed multiple testing criteria highlighted in green. (C) Miami plot comparing postpartum hemorrhage due to atony (top) and retained placenta (bottom). Green dots indicate SNPs passing the multiple testing threshold.

**Figure 2.**
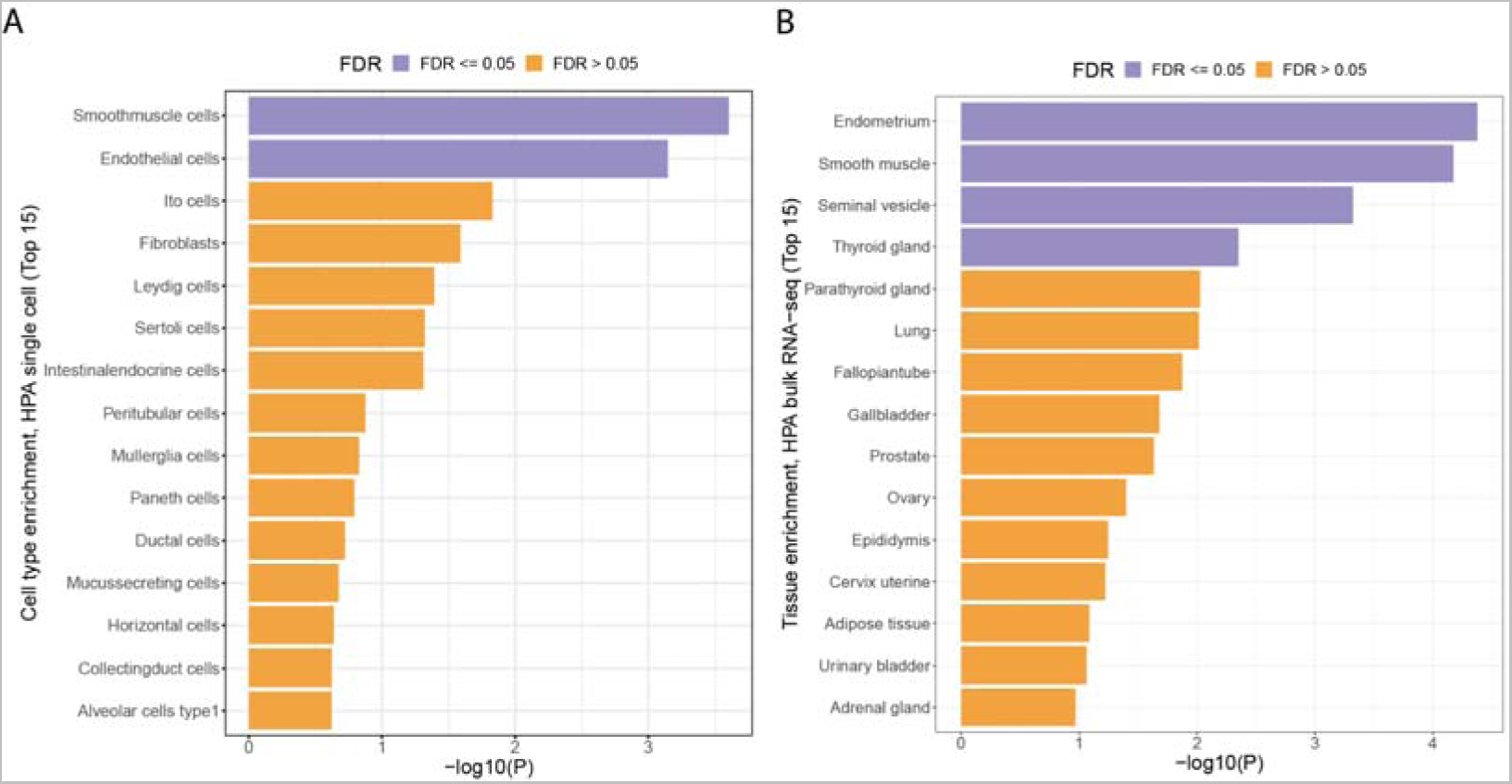
(A) MAGMA single cell enrichment from the Human Protein Atlas. Smooth muscle cells and endothelial cells were both enriched (FDR < 0.05) (C) MAGMA bulk tissue enrichment from the Human Protein Atlas showed an enrichment of endometrial, smooth muscle, seminal vesicle, and thyroid gland tissue (FDR < 0.05).

**Table 1.**
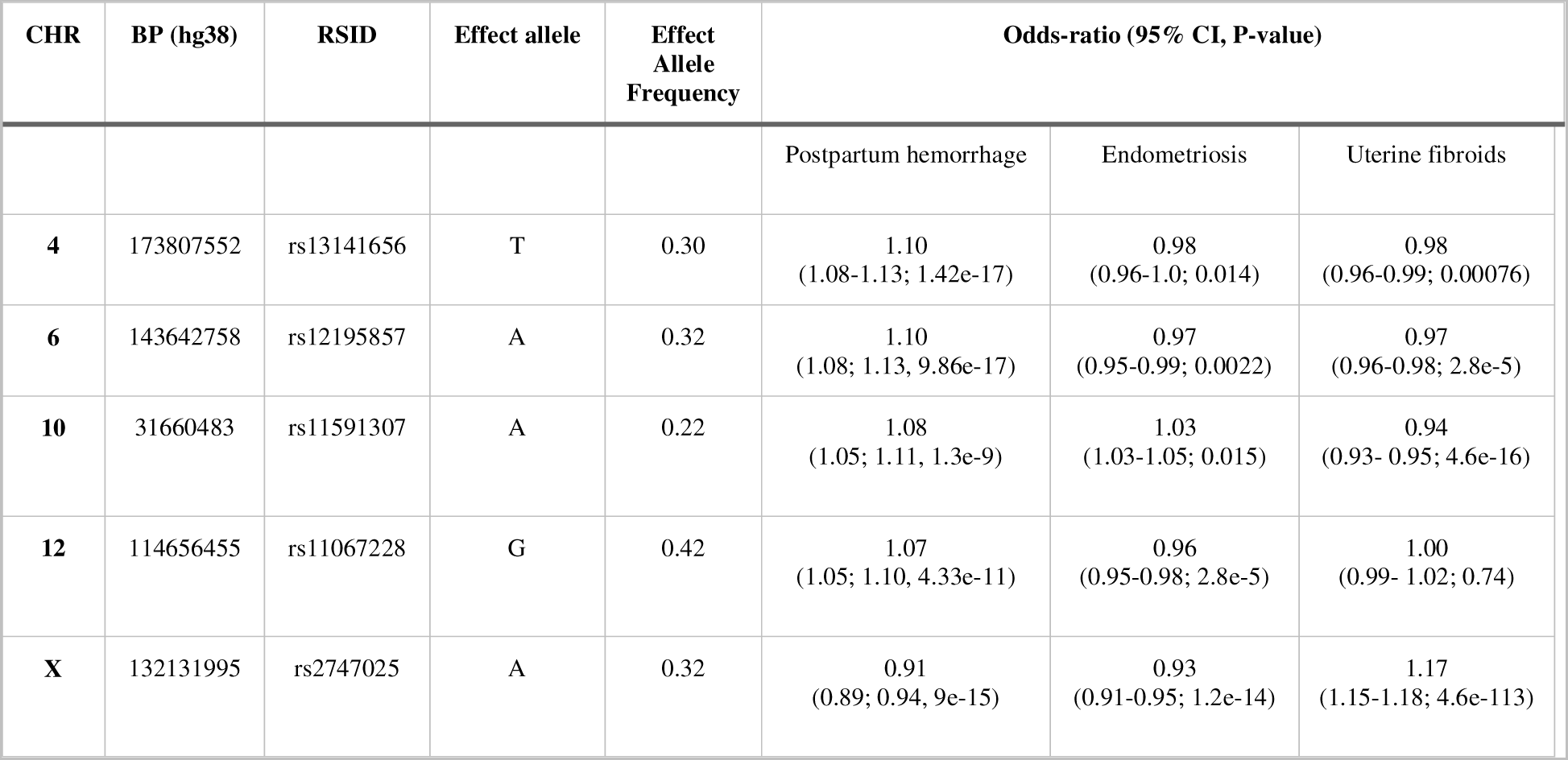
Effect sizes across loci for PPH, endometriosis and uterine fibroids. Endometriosis and uterine fibroid estimates come from the datasets listed in Supplementary Table 4.

### Prior evidence of SNPs

According to the GWAS catalog^10^, the lead variant on chromosome 12 has previously been found in association with heel bone mineral density and prostate-specific antigen levels in males, both of which hormone-responsive tissues. Additionally, the lead variants on chromosomes 10 and X were in strong (*r*^2^ >0.8) linkage disequilibrium (LD) with variants associated with uterine fibroids and endometriosis, while the lead variant on chromosome 6 was in strong LD with a sequence variant associated with educational attainment (Supplementary Table 4). Furthermore, we investigated the genome-wide significant lead variants in the FinnGen cohort (R9) and found that the lead variants on chromosome 12 (*TBX3*) and chromosome X (FRDM7/*RAP2C*) were also associated with endometriosis, and the loci on chromosomes 6 (*PHACTR2*), 10 (*ZEB1*), and X (FRDM7/*RAP2C*) were associated with uterine fibroids (Table 2) (Supplementary Figure 1B).

**Table 2.** PPH signals were enriched (p <0.05, Bonferroni corrected) within binding sites for progesterone receptor (PGR) defined in human embryonic stem cells (hESC). Shown are nominally significant results i.e., where uncorrected p-value <0.05. We defined binding sites by ChIP-seq data available through Remap2022 database (website: remap.univ-amu.fr).

| DNA binding protein | Tissue /Cell line | Annotated PPH signals, n | Expected proportion of annotated PPH signals | $p$-value |
| --- | --- | --- | --- | --- |
| PGR | hESC | 4/5 | 5% | 5e-06 |
| ZNF558 | HEK293 | 4/5 | 27% | 7e-05 |
| PGR | myometrium | 5/5 | 17% | 0.001 |
| PGR | leiomyoma | 3/5 | 8,6% | 0.002 |
| IRF2BP2 | HEK293 | 3/5 | 9,9% | 0.005 |
| MED12 | leiomyoma | 3/5 | 11% | 0.007 |
| MYOG | RH4 | 3/5 | 11% | 0.009 |
| MED12 | myometrium | 3/5 | 14% | 0.01 |
| ONECUT1 | Hep-G2 | 3/5 | 15% | 0.019 |
| FOXA1 | prostate | 4/5 | 17% | 0.02 |
| ZNF3 | Hep-G2 | 3/5 | 7% | 0.023 |

### Functional analysis of loci

We annotated the five PPH lead variants and their correlated variants (r^2^>0.80), hereafter referred to as PPH signals, according to their location in the ENCODE encyclopedia of candidate cis-regulatory elements (cCRE)^11^. Collectively, cCREs span 291Mb of the genome and contain 10.2% of sequence variants. We found that all five PPH signals were located within either the distal or proximal enhancer-like sequences (dELS, pELS), suggesting non-coding regulatory functions (Supplementary Table 55-7).

The predicted gene targets for these regulatory elements in uterine tissue are *TBX3* (12q24.21), *FRMD7* and *RAP2C* (Xq26.2) according to Epimap^12^ (Supplementary Table 8-8). Furthermore, there is evidence that the lead SNP at the chromosome 4 locus, rs13141656, targets *HAND2* in endometrial tissue^13, 14^. None of these genes have been directly associated with PPH. *HAND2* and *TBX3* are involved in stromal-epithelial communication during implantation. *HAND2* is implicated in preterm birth and gestational duration and has previously been found to be critical for implantation^15, 16^. The function of the *RAP2C/FRMD7* gene cluster is currently unknown, but variants in the *RAP2C* locus are associated with gestational duration^17^. None of the proteins are known to physically interact, according to the STRING database (v11.5)^18^.

We tested the PPH signals for enrichment within 1,210 transcription factor (TF) binding sites in DNA of various cell types and tissues^19^, amounting to a total of 4,143 tests and we used Bonferroni correction to set the threshold for significances at p<0.05/4,143 ∼ 1·10^-5^. The number of PPH signals found in PGR binding sites in human embryonic stem cells was significantly higher than expected (p=5·10^-6^, Table 2). PGR is an important factor in the establishment and maintenance of pregnancy and is therefore relevant in the context of PPH.

We used MAGMA^20^ to test for tissue-specific enrichment using expression data from the Human Protein Atlas bulk tissue and single-cell datasets^21^. We found that the endometrium, smooth muscle, seminal vesicle, and thyroid gland tissue were enriched, as well as endothelial cells (FDR < 5%) (Figure 2A,B).

### Maternal and fetal transmission

We performed a haplotype-specific analysis of the five PPH-associated variants in the MoBa and deCODE cohorts to distinguish between maternal and fetal effects. These results were consistent with all five variants affecting the risk of PPH primarily through the maternal genome (Supplementary Figure 5, Supplementary Table 10). However, we cannot exclude any effect from the fetal genome.

### Heritability of pregnancy-associated bleeding traits

We estimated the SNP heritability of early bleeding in pregnancy and PPH to be 12.7% (95% CI 7.8-17.6%) and 16.5% (95% CI 10.2-22.8%), respectively, in the Danish cohort, assuming a population prevalence of 25% and 15%, respectively. We selected prevalence’s based on literature review^2, 8^.

#### Genetic correlations between pregnancy-associated bleeding traits

We characterized the intra-phenotypic genetic correlations among the five bleeding in pregnancy phenotypes investigated in this study: “early bleeding in pregnancy, any outcome”, “early bleeding in pregnancy, live birth”, “PPH”, “PPH due to atony”, and “PPH due to retained placenta”. Antepartum hemorrhage did not have sufficient polygenic signal to be investigated (LDSC x^z^ < 1.02). Early bleeding during pregnancy did not exhibit any significant genetic correlation with PPH or any of its subtypes (Figure 3A). Notably, there was strong genetic correlation between PPH due to uterine atony and PPH due to retained placenta (r_g_=0.77, 0.49-1.05 95% CI).

**Figure 3.**
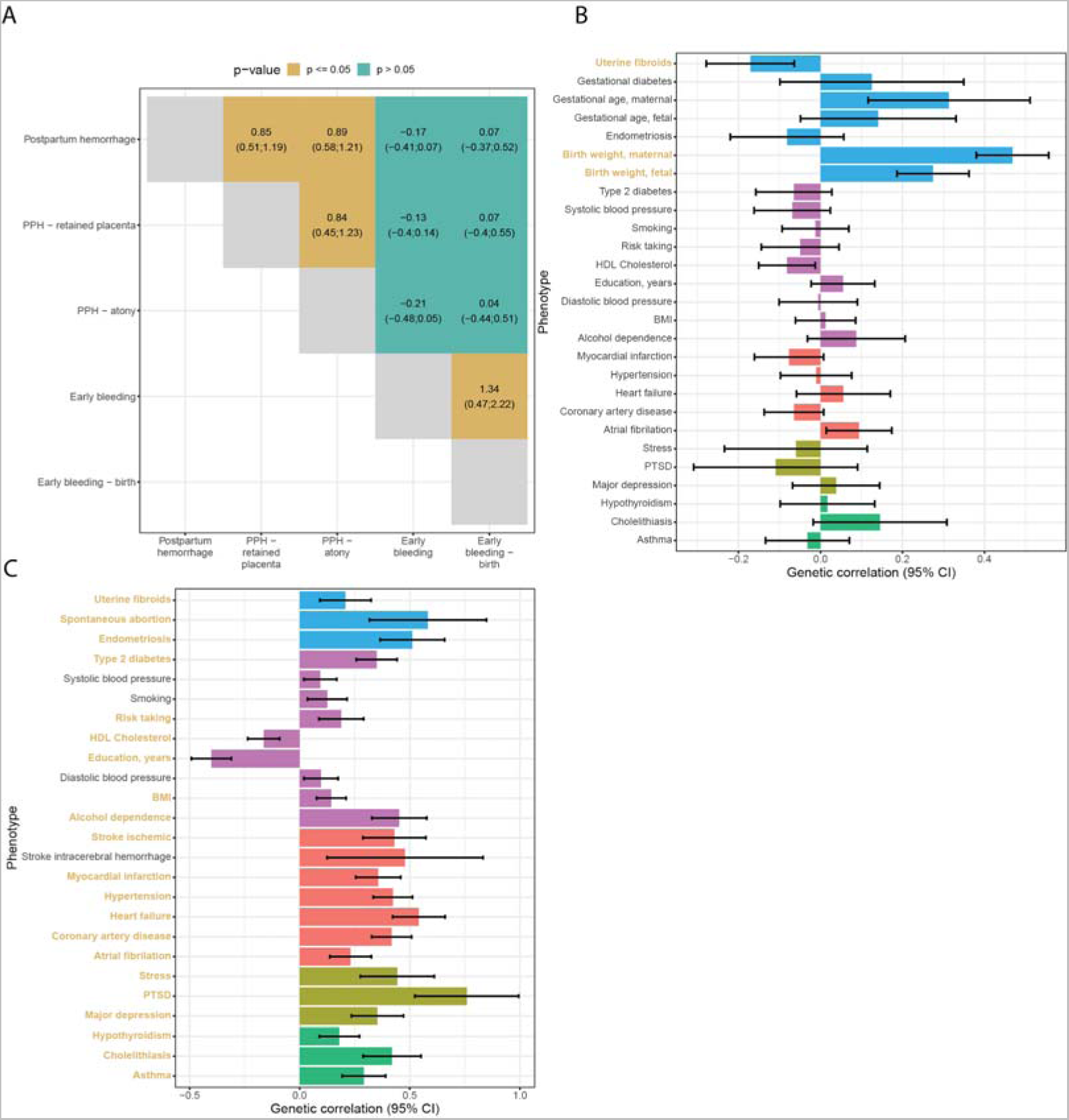
(A) Cross-trait genetic correlation of all bleeding in pregnancy phenotypes (95% confidence interval). Postpartum hemorrhage and early bleeding in pregnancy show no noteworthy genetic correlation. Postpartum hemorrhage due to atony or retained placenta are genetically indistinguishable. (B) Genetic correlations between postpartum bleeding and selected disorders. (C) Genetic correlations between early bleeding and selected traits. Correlations that are significant after accounting for the number of traits tested are highlighted in yellow. Error bars represent the 95% confidence interval. The data sets used for the analysis are described in Supplementary Table 5.

#### Phenotypes correlated with pregnancy-associated bleeding

Next, we aimed to characterize the genetic overlap of early bleeding (any outcome) and PPH with other co-occurring diseases and other phenotypes. The range of phenotypes that may co-occur with early bleeding during pregnancy and PPH has not been extensively characterized. Consequently, we looked for associations in three distinct cohorts: the Estonian Biobank (n=17,094), UK Biobank (n=12,490), and a Danish nationwide cohort (n=2,320,776). Following a meta-analysis of 417 and 628 ICD-10 codes at the third level for early bleeding and PPH, respectively, we found that 120 codes were significantly associated with PPH (FDR < 0.05) and 625 codes with early bleeding (Supplementary File 1).

Based on the literature, known risk factors, lifestyle, socioeconomic factors, and the pairwise phenotype-to-phenotype correlation analyses presented here, we identified a list of phenotypes for which we could find suitable summary statistics (Supplementary Table 11). We additionally included socioeconomic and cardiometabolic traits, such as BMI, smoking, and blood pressure. These traits are not recoded in the registries, but are highly correlated with the diseases we found in the phenotype-to-phenotype correlation analysis. PPH was, at the genetic level, strongly positively correlated with birth weight (maternal and fetal), gestational duration (maternal), and had an inverse correlation with uterine fibroids (Bonferroni-corrected *p* < 0.05) (Figure 3B, see Supplementary Table 11 for a description of the summary stats). No other traits displayed a significant genetic correlation with PPH after multiple testing corrections. Although no sequence variants were found in association with early bleeding, we nonetheless, found genetic correlations to reproductive, socioeconomic, cardiovascular, and psychiatric traits (Figure 3C).

### Polygenic risk scores

Utilizing 25,118 pregnancies (n=19,026 women) since 2012 from the Danish cohort, we found that a logistic regression model including the polygenic risk score (PRS) for PPH and birth weight yielded an improved model (p < 2 · 10^-16^, likelihood ratio test, Supplementary Table 12), compared to a model that included only age, pre-pregnancy BMI, parity, prior number of cesarean sections, and prior number of PPHs. The variance explained (Nagelkerke R^2^) increased from 3.2% (2.7%; 3.8%) to 3.8% (3.4%; 4.5%), yielding a net improvement of 0.7% (0.5%; 0.9%). Similarly, the AUC increased from 0.60 (0.59; 0.61) to 0.61 (0.60; 0.62), improving marginally (0.008, 0.005; 0.011).

## Discussion

### Summary

In this study, we investigated the genetic architecture of bleeding associated with pregnancy, which is one of the most common complications of pregnancy associated with both maternal and fetal morbidity and mortality. We identified five loci associated with PPH, with strong functional evidence of association with genes involved in implantation and contraction. Furthermore, enrichment of progesterone receptor binding sites substantiates the importance of hormone regulation in the etiology of PPH and suggests organ-specific dysregulation. However, in the absence of relevant tissue (myometrium sampled during or right before pregnancy), we were not able to locate the point or points in pregnancy at which the sequence variants exert their effect. There was no evidence of a genetic correlation between PPH and diseases. Our study revealed that early bleeding is highly polygenic with genetic correlations spanning various different categories of human traits, and PPH is a disorder of hormone-responsive genes. Overall, this study provides new insights into the genetic basis of bleeding during pregnancy, and suggests different genetic pathways for early bleeding and PPH.

### Strengths and limitations

In this study, data from six Northern European cohorts were analyzed, representing six different countries with similar, albeit varying, universal healthcare systems, protocols for pregnancy care, and levels of available clinical information. However, it is important to note that PPH disproportionately affects women in developing countries, and further research is needed to integrate more diverse populations into studies of this kind. Additionally, the registration of early bleeding during pregnancy depends heavily on the healthcare-seeking behavior of the individual, organization of early pregnancy care and is most likely affected by the heterogeneous causes of early bleeding. Not all cohorts had information on early bleeding during pregnancy, and only three cohorts could distinguish between events leading to live births and those that did not. Another factor that should be considered is that oxytocin, a drug used to prevent or treat PPH, is administered preemptively based on other factors, such as cesarean section and PPH in a previous pregnancy. This bias most likely results in a smaller effect, thereby requiring a larger sample size for detection of associated loci.

### Comparison with other literature

In this study, the potential causal genes at the five loci that may contribute to the development of PPH were not related to previously suggested causes, such as the oxytocin receptor or coagulation cascade^5, 22^. The latter being expected as women with known coagulation disorders were excluded. The identified loci were found to be significantly enriched with progesterone-binding sites in human embryonic stem cells and showed nominal significance in the myometrium, the smooth muscle layer of the uterus responsible for contractions during labor and delivery. Progesterone is known to relax the myometrium and reduce contractility^23^, which is vital for maintaining a healthy pregnancy. The presence of progesterone-binding sites suggests that the genes located in these regions may be involved in regulating myometrial contractility, and that abnormal contractions can lead to PPH. Furthermore, these loci were also associated with endometriosis and/or uterine fibroids. Endometriosis and uterine fibroids are both treated with Selective Progesterone Receptor Modulators (SPRM), which target the progesterone receptor^24^. Observational studies suggest that early bleeding, antepartum hemorrhage, and postpartum hemorrhage are correlated^2, 25^. However, we did not observe any evidence of a shared genetic etiology.

We established early bleeding as a complex trait, substantiated by significant heritability, polygenic signals, and widespread pleiotropy across disease areas. Early bleeding is related to pregnancy loss and may be an indication of the maternal body not coping well with the pregnancy. Genetic correlation with post-traumatic stress disorder and a variety of seemingly unrelated diseases and traits may be an indication of an extreme response to stress and a general low tolerance of the added burden of pregnancy upon maternal systems with underlying weaknesses. Possibly due to the high heterogeneity in the phenotype, we did not identify any variants associating with early bleeding; therefore, we could not test for causality using e.g., Mendelian randomization. Nonetheless, a previous study indicated a causal relationship between early bleeding and cardiometabolic diseases^1^.

The use of polygenic risk scores resulted in marginal improvements in the predictive capability for PPH. Nonetheless, as genetic studies become better powered, we can expect an improvement in their predictive capability. Consequently, the addition of polygenic risk scores to prognostic models should be considered in future studies to enable early stratification of women at a high risk of PPH.

## Conclusion

Our findings reveal complex genetics of early bleeding in pregnancy. They further provide valuable insights into the potential underlying mechanisms of PPH and may inform the development of more effective prevention strategies.

## Methods

### Study Cohorts

This was a multi-national study that included six cohorts of Western European ancestry: the Copenhagen Hospital Biobank study on Reproduction (Denmark), Estonian Biobank (Estonia), FinnGen (Finland), deCODE genetics (Iceland), UK Biobank (England), and Norwegian Mother, Father and Child Cohort Study (Norway). All studies were approved by the relevant institutional ethics review boards (Supplementary Text)

### Copenhagen Hospital Biobank study on Reproduction and the Danish Blood Donor Study

The Copenhagen Hospital Biobank (CHB) is based on EDTA blood samples collected from patients for blood typing and red cell antibody screening at hospitals in the Greater Copenhagen Area^26^. The CHB study on Reproduction (CHB-Repro) cohort focuses on patients with fertility and obstetric complications, identified through the Danish National Patient Registry. We also included blood donors from the Danish Blood Donor Study Genomic Cohort (DBDS-GC). DBDS-GC is described by Hansen et al^27^. All samples were genotyped at deCODE genetics using the Illumina Infinium Global Screening array. Samples were imputed using an in-house pan-Scandinavian reference panel^28^. Association analysis was performed using software developed at deCODE genetics^29^.

### Estonian Biobank

The EstBB is a population-based biobank with over 200,000 participants (corresponding to 20% of the total Estonian population). Details of EstBB genotyping procedure have been described previously^30, 31^. Briefly, all EstBB participants were genotyped using Illumina arrays at the Core Genotyping Lab of the Institute of Genomics, University of Tartu. Samples were imputed using a population specific imputation reference of 2,297 whole genome sequencing samples^32^. Association analysis was performed using SAIGE 0.43.1.

### FinnGen

FinnGen is a public–private partnership research project that combines imputed genotype data generated from newly collected and legacy samples from Finnish biobanks and digital health record data from Finnish health registries (https://www.finngen.fi/en) with the aim to provide new insights into disease genetics^33^. FinnGen includes 9 Finnish biobanks, research institutes, universities and university hospitals, 13 international pharmaceutical industry partners and the Finnish Biobank Cooperative (FINBB) in a pre-competitive partnership. As of November 2022 (release 10 described in this article), samples from 412,181 individuals have been analysed with the final aim to have a cohort of 500,000 participants. The project utilizes data from the nationwide longitudinal health register collected since 1969 from every resident in Finland.

### deCODE genetics

The deCODE cohort is a nation-wide sample collection recruited in Iceland since 1997. All participants who donated blood signed an informed consent. Variants were identified through whole genome sequencing of 63,460 individuals. They were imputed into 173,025 chip-genotyped Icelanders using long-range phasing, and into their untyped close relatives based on genealogy^29, 34^. We used logistic regression to test for association of sequence variants assuming an additive genetic model, using software developed at deCODE genetics^29^.

### Norwegian Mother, Father and Child Cohort Study

The Norwegian Mother, Father and Child Cohort Study (MoBa) is a population-based pregnancy cohort study conducted by the Norwegian Institute of Public Health. Participants were recruited from all over Norway from 1999-2008^35^. The women consented to participation in 41% of the pregnancies. The cohort includes approximately 114.500 children, 95.200 mothers and 75.200 fathers. The current study is based on version 12 of the quality-assured data files released for research. Details about PPH were obtained from the Medical Birth Registry, a national health registry containing information about all births in Norway. Sample QC and imputation has previously been described^36^. In brief, individuals were genotyped using different Illumina arrays (HumanCoreExome-12 v1.1, HumanCoreExome-24 v1.0, Global Screening Array v1.0, InfiniumOmniExpress-24 v.2, HumanOmniExpress-24 v1.0). Individual level QC was performed to remove ancestry outliers and individuals with sex discrepancy and call rate < 0.98. Furthermore, SNPs with a MAF < 1%, deviating from the Hardy-Weinberg equilibrium (p < 1e-4), or a call rate < 0.98 were removed. Imputation was done using SHAPEITv2 + PBWT on the Sanger imputation server, with HRC v1.1 as the imputation reference panel. Association analysis was done using regenie^37^.

### UK Biobank

The UK Biobank is a prospective cohort of ∼500.000 individuals from across the United Kingdom, recruited at ages 40-69. Genotyping was done in two batches, using the Affymetrix chip UK BiLEVE Axiom87 and Affymetrix UK Biobank Axiom array. Imputation was done using a sample of 150,000 whole genome sequenced individuals from the UK Biobank^38^. Only individuals with a registered live or stillbirth (identified through the HESIN delivery table) and of European descent were included in the analysis. Association analysis was performed using software developed at deCODE genetics^29^. The UKB resource was used under application no. 56270. All phenotype and genotype data were collected following an informed consent obtained from all participants.

### Phenotype definitions

We divided bleeding in pregnancy into three categories and the following sub-phenotypes:

1. Bleeding in early pregnancy (<20+0 gestational weeks)

a. Bleeding in early pregnancy leading to live birth
b. Bleeding in early pregnancy ending in any outcome (live birth, pregnancy loss, termination of pregnancy, ectopic pregnancy, mola pregnancy, pregnancy of unknown location)
2. Antepartum hemorrhage (>20th gestational week, prior to birth)
3. Postpartum hemorrhage (PPH, hemorrhage following birth)

a. PPH due to atony
b. PPH due to retained placenta

We categorized each phenotype using hospital admission codes, although not all codes were available in all countries. We provided a phenotype definition list in Supplementary Table 13. We adjusted analyses for age, parity, gestational duration, and weight of the child, if possible. Women with known coagulation disorders were excluded (ICD-10 codes D66-D69, O46.0, O67.0). Furthermore, we excluded multifold pregnancies for antepartum hemorrhage and PPH, if possible. Lastly, we excluded pregnancies delivered by cesarean section in the PPH analysis, if possible.

### Meta-analysis

For the meta-analyses, we combined GWASs from the respective cohorts using a fixed-effects inverse variance method based on effect estimates and standard errors in which each dataset was assumed to have a common odds-ratio but allowed to have different population frequencies for alleles and genotypes. Sequence variants were mapped to NCBI Build38 and matched on position and alleles to harmonize the datasets. After excluding variants with discrepant allele frequency between cohorts, variants with MAF < 0.001% in all cohorts or variants only present in one dataset, 18,009,056 variants were included in the meta-analysis. The threshold for genome-wide significance was corrected for multiple testing with a weighted Bonferroni adjustment that controls for the family-wise error rate, using as weights the enrichment of variant classes with predicted functional impact among association signals^39^. The significance threshold then becomes 4.56 × 10^-7^ for high-impact variants (including stop-gained, frameshift, splice acceptor or donor), 9.12 × 10^-8^ for moderate-impact variants (including missense, splice-region variants and in-frame indels), 8.28 × 10^-9^ for low-impact variants (synonymous, 5’ and 3’ UTR, upstream and downstream variants), 4.19 × 10^-9^ for other DNase I hypersensitivity sites (DHS) variants and 1.38 × 10^-9^ for other non-DHS variants. In a random-effects method, a likelihood ratio test was performed in all genome-wide associations to test the heterogeneity of the effect estimate in the four datasets; the null hypothesis is that the effects are the same in all datasets and the alternative hypothesis is that the effects differ between datasets.

### Conditional analysis

Conditional association analyses were performed on the GWASs from Iceland, the UK, and Denmark using true imputed genotypes of participants. Approximate conditional analyses (COJO), implemented in the GCTA-software, were applied on the lead variants in the Finnish, Estonian and MoBa summary statistics^40, 41^. Linkage disequilibrium between variants was estimated using a set of 5,000 WGS Icelanders. The analyses were restricted to variants within 1 Mb from the index variants. The p-values were combined for all six datasets to identify any secondary signals. Based on the number of variants tested we required secondary signals to pass a threshold of p < 5 × 10^-8^ after correcting for the lead variant.

### Comparison of effect sizes for retained placenta and uterine atony

We compared effect sizes for retained placenta and uterine atony by doing a case-case analysis of the summary statistics using ReAct^42^. Only genome-wide significant SNPs, according to the functionally informed multiple testing correction, found in the main analysis of PPH was included. We assumed no overlap between cases, and a full overlap between controls.

### Lookup of variants

Variants and variants in strong LD were looked up in the GWAS catalog to identify prior associations to other phenotypes, using the LDlinkR package^10, 43^. Furthermore, we investigated the association of the variants to endometriosis and uterine fibroids in the FinnGen cohort (r10). The analysis was part of the FinnGen core analysis, done using regenie, in which the analysis was adjusted for age, the first ten principal components, genotyping chip, and batch^37^. We adjusted p-values for the number of phenotypes (two) and variants (234) tested (p<0.05/(2 · 234) = 0.0001).

### Mapping of GWA signals to non-coding annotations

We downloaded annotations of candidate cis-regulatory elements (cCRE; version 3) from the ENCODE project (*website: screen.encodeproject.org*)^10^. We then determined whether the lead PPH sequence variant or any of their correlated variants (*r*^2^ > 0.80), i.e. PPH signals were located within cell-type agnostic cCREs (candidate cis-regulatory elements), and cCREs defined in tissue samples relevant to PPH i.e. uterus tissue. In this same way, we annotated the PPH signals with respect to enhancer elements (Active/Genic) as defined for 833 samples (representing 33 groups of tissues/organs) in EpiMap (*website: compbio.mit.edu/epimap*)^12^. EpiMap further provides predicted links between enhancers and genes, and, based on these pre-computed predictions, we looked for candidate gene targets for each signal in uterus tissue (*website: personal.broadinstitute.org/cboix/epimap/links/links_corr_only*). We also annotated the PPH signals with respect to DNA binding sites for 1,210 transcription factors (TFs) mapped experimentally by various researchers, notably ENCODE project, using ChIP-seq in different tissue/cell types and conditions made available by Remap2022 (*website: remap2022.univ-amu.fr*), which amount to a total of 4,143 ChIP-seq experiments.

### Enrichment of association signals in functional annotations

We used GWA signals from the GWAS catalog (see details in next paragraph: „*GWAS catalog*“) to obtain the null distribution in our enrichment analyses for functional annotations of the genome. The number of sequence variants found in high linkage disequilibrium (LD; *r*^2^>0.80) for each of the five PPH association signals were expected to influence the probability of finding an overlap to a given functional annotation map. We therefore randomly selected five GWA signals from the GWAS catalog for each of the five PPH signals, ensuring that the five randomly selected signals were matched to the PPH signals with respect to the number of sequence variants found in high LD. We then counted the number of randomly selected signals that intersected with a given annotation (this count is denoted as z). This procedure was then repeated N = 200,000 times. In summary, we were simulating the five PPH signals in terms of a) the number of sequence variants in high LD to each PPH signal and b) the property of being a GWA signal associated with human multifactorial trait.

Let *z_i_* represent the number of annotated signals in each *i*-th sample. The probability (p) of finding an intersection to a given annotation among randomly sampled GWA signals is therefore: 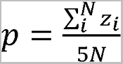, where 5 *N* = the total number of randomly sampled GWA signals from the GWAS catalog (five randomly selected GWA signals in each of *N* samples); this is the expected proportion of annotated GWA signals. We then define *X* ∼ *Bin*(*n, p*) where *X* is the number of annotated PPH signals and n is the number of PPH signals (*n* = 5). The five PPH signals are found on different chromosomes, and we therefore assume that they are independent. We then determine the probability of observing *x* or more PPH signals in a given annotation, where *x* is the observed number of PPH signals that intersect with the given annotation. We are therefore interested in: (*X* ≥ *x*) = *j/*N, where *j* is the number of times we found *x* or more annotated GWA signals in the aforementioned *N* random samples of GWA signals. We then used Bonferroni correction to set the threshold for significance.

*GWAS catalog*: We compiled a robust set of association signals from the NHGRI-EBI catalog of GWAS association signals; downloaded on 4-AUG-2021 (GWAS catalog v1.00 website: www.ebi.ac.uk/gwas)^10^. GWAS catalog variants (lead) were matched to in-house variant calls on the basis of rs-identifiers, genome position and MAF (GWAS catalog entries with missing information in any of these fields were omitted). In the GWAS catalog, the same trait has been studied by many different research groups and therefore many associations are „repeated“ and therefore not independent. We used the following procedure to compile a set of independent associations for each trait in the GWAS catalog: First, we extracted all associations with the trait with *P*-value<1e-9. Second, we selected the most significant association and added it to the list of independent associations. Third, we added the most significant associations with *P*-value<1e-9 located more than 1Mb away from other independent associations. We then repeated this third step until no more associations were found with *P*-value<1E-9 while also located >1Mb away from those already added to the list of independent associations. We omitted traits classified as „blood protein measurement“ (mostly representing GWASs for serum protein assays) and sixteen other traits (e.g. heel bone mineral density) with an unusually large number of associations. Further, as our enrichment method takes LD into account (computed in whole genome sequenced individuals from the Icelandic population), we selected GWAŚs carried out in individuals of European descent. This resulted in 27,546 GWA association signals for 1,173 diseases or other human traits.

### Functional enrichment and tissue specificity

We used MAGMA to investigate tissue expression specificity^20^. Consensus bulk and single-cell RNA-Seq data that had already been preprocessed was downloaded from the Human Protein Atlas^21^. In short, the HPA consensus tissue gene data summarizes expression at the gene level covering 62 tissues, and includes data from the Human Protein Atlas, GTEx, and FANTOM5. The RNA single cell consensus data set covers 51 cell types across 13 tissues, from 14 different studies. We used the 1,000 Genomes Phase 3 European data as reference (downloaded from https://ctg.cncr.nl/software/magma).

### Comorbidity analysis

Comorbidities associated with early bleeding in pregnancy and PPH were identified across three cohorts (Denmark, Estonian Biobank, and the UK Biobank). The Danish cohort utilized nationwide data from the Danish National Patient Register (DNPR) and the Danish Medical Birth Register (DMBR)^44, 45^. The DNPR contains hospital admissions since 1977, and the DMBR contains birth since 1973. We identified all women born after 1957, which ensured a full reproductive history from their 20th year birthday and onwards. We analyzed associations between early bleeding in pregnancy, PPH and all other diagnoses (excluding chapters regarding infections, obstetric diagnosis, injuries, and contacts with the healthcare system). Similarly, a PheWAS was performed in the Estonian Biobank and the UK Biobank. In the UK Biobank, we included only women present in the HESIN delivery tables. Odds ratios were determined using logistic regression, adjusting for year of birth. Data from the three cohorts were meta-analyzed using an inverse-variance weighting as implemented on the R package metafor. We controlled for multiple testing by calculating q-values and selecting associations with a q-value < 0.05.

### Heritability and genetic correlations

SNP Heritability was estimated using RHE-mc, which is an efficient and scalable estimator using individual level data^46^. We selected genotyped SNPs in the CHB with MAF > 1%, missing in less than 1% of samples, no deviation from HWE (p < 10^-7^), and excluded the HMC region, as per author’s recommendations. We adjusted the analysis for year of birth, year of birth squared, and the first 10 principal components.

Genetic correlations were estimated using LD Score Regression^47^. We selected phenotypes based on prior knowledge about risk factors and associations from the comorbidity analysis and availability. In this analysis, we used results for about 1.2 million well imputed variants, and for LD information we used precomputed LD scores for European populations (downloaded from: https://data.broadinstitute.org/alkesgroup/LDSCORE/eur_w_ld_chr.tar.bz2). Genetic correlation of pregnancy bleeding subtypes was calculated between Danish primary trait and the meta-analysis of the relevant secondary trait, excluding Danes, and vice versa. The results of the two analyses were then meta-analyzed. Genetic correlation of Early bleeding - birth was only done using the Danish data for the primary trait as the sample size for the remaining populations was too small.

### Polygenic Risk Scores

Polygenic Risk Scores (PRS) were created using LDPred2^48^. Autosomal genotype data from 138,669 individuals in the Copenhagen Hospital Biobank study on Reproduction was filtered to only include variants present in LDpred2’s recommended set of 1,054,330 reference variants. Missing genotype information was imputed to be the affected locus’ reference allele. GWAS Summary statistics for birth weight from Warrington et al was pre-processed with MungeSumStats^49, 50^. The birth weight summary statistics contain a very small fraction of Danish samples from other cohorts. We excluded any Danes from the summary statistics used for the PPH PRS to avoid inflation.

The effects of polygenic risk scores were estimated using a logistic regression model, adjusted for maternal age at conception, parity, pre-pregnancy BMI, previous number of cesarean sections, and previous numbers of PPH events. We compared models with and without polygenic risk scores using a likelihood ratio test. Furthermore, we also compared the C-index and Nagelkerke’s R^2^. We used a bootstrap resampling approach to find optimism corrected values, which is a conservative estimate of the error on unseen data and a method of performing an internal validation^51^. We repeated the bootstrap resampling 100 times, and we report the 95% percentile bootstrap confidence intervals. Standard errors were corrected for the inherent clustering present due to multiple pregnancies from the same women using the Huber-White method.

### Haplotype analysis

We explored whether the effects of the identified variants on PPH depend on maternal, fetal or maternal and fetal origins by performing an association analysis using the parental transmitted and non-transmitted alleles. We used phased genotype data from the MoBa cohort (n = 22,330 parent-offspring trios) and deCODE study to infer the parent of origin of fetal alleles. The analysis of the deCODE data was done on 106,622 parent-offspring trios (2,558 cases and 104,064 controls) with at least one genotyped individual. This included 19,488 fully genotyped trios, 5,991 with only child and mother and 1,835 with only child and father genotyped, 39,390 with both parents genotyped but not the child, and 1,661, 26,582 and 11,675 with only child, mother or father genotyped, respectively.

For each lead variant, the following logistic regression model was fit:

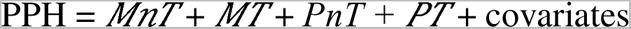

where *MnT* and *MT* refer to the maternal non transmitted and transmitted alleles, respectively, and *PnT* and *PT* refer to the paternal non-transmitted and transmitted alleles, respectively. The *PT* effect is interpreted as a fetal-only genetic effect, whereas the effect of the maternal non transmitted allele is a maternal-only genetic effect. In the deCODE study, we used maximum likelihood estimation to estimate the effects, as previously described^52^. Estimates from the two cohorts were meta-analyzed using fixed-effect meta-analysis.

### Data availability

Meta-analysis summary statistics will be made available upon publication.

## Acknowledgements

The work is carried out as a part of the BRIDGE – Translational Excellence Programme (bridge.ku.dk) at the Faculty of Health and Medical Sciences, University of Copenhagen, funded by the Novo Nordisk Foundation. Grant agreements NNF18SA0034956, NNF14CC0001, and NNF17OC0027594. Furthermore, we would like to acknowledge funding from the Ole Kirk Foundation and Rigshospitalet’s Research Fund.

B.J. received funding from The Swedish Research Council, Stockholm, Sweden (2019-01004), The Research Council of Norway, Oslo, Norway (FRIMEDBIO #547711), March of Dimes (#21-FY16-121), Agreement concerning research and education of doctors (ALFGBG-965353). Research by B.J. was also supported by the Eunice Kennedy Shriver National Institute Of Child Health & Human Development of the National Institutes of Health under Award Number R01HD101669. The content is solely the responsibility of the authors and does not necessarily represent the official views of the National Institutes of Health. We thank the Norwegian Institute of Public Health (NIPH) for generating high-quality genomic data. This research is part of the HARVEST collaboration, supported by the Research Council of Norway (#229624). We also thank deCODE genetics and the NORMENT Centre for providing genotype data, funded by the Research Council of Norway (#223273), South East Norway Health Authorities and Stiftelsen Kristian Gerhard Jebsen. We further thank the Center for Diabetes Research, the University of Bergen for providing genotype data and performing quality control and imputation of the data funded by the ERC AdG project SELECTionPREDISPOSED, Stiftelsen Kristian Gerhard Jebsen, Trond Mohn Foundation, the Research Council of Norway, the Novo Nordisk Foundation, the University of Bergen, and the Western Norway Health Authorities.

We want to acknowledge the participants and investigators of FinnGen study. The FinnGen project is funded by two grants from Business Finland (HUS 4685/31/2016 and UH 4386/31/2016) and the following industry partners: AbbVie Inc., AstraZeneca UK Ltd, Biogen MA Inc., Bristol Myers Squibb (and Celgene Corporation & Celgene International II Sàrl), Genentech Inc., Merck Sharp & Dohme LCC, Pfizer Inc., GlaxoSmithKline Intellectual Property Development Ltd., Sanofi US Services Inc., Maze Therapeutics Inc., Janssen Biotech Inc, Novartis AG, and Boehringer Ingelheim International GmbH. Following biobanks are acknowledged for delivering biobank samples to FinnGen: Auria Biobank (www.auria.fi/biopankki), THL Biobank (www.thl.fi/biobank), Helsinki Biobank (www.helsinginbiopankki.fi), Biobank Borealis of Northern Finland (https://www.ppshp.fi/Tutkimus-ja-opetus/Biopankki/Pages/Biobank-Borealis-briefly-in-English.aspx), Finnish Clinical Biobank Tampere (www.tays.fi/en-US/Research_and_development/Finnish_Clinical_Biobank_Tampere), Biobank of Eastern Finland (www.ita-suomenbiopankki.fi/en), Central Finland Biobank (www.ksshp.fi/fi-FI/Potilaalle/Biopankki), Finnish Red Cross Blood Service Biobank (www.veripalvelu.fi/verenluovutus/biopankkitoiminta), Terveystalo Biobank (www.terveystalo.com/fi/Yritystietoa/Terveystalo-Biopankki/Biopankki/) and Arctic Biobank (https://www.oulu.fi/en/university/faculties-and-units/faculty-medicine/northern-finland-birth-cohorts-and-arctic-biobank). All Finnish Biobanks are members of <u>BBMRI.fi</u> infrastructure (www.bbmri.fi). Finnish Biobank Cooperative -FINBB (https://finbb.fi/) is the coordinator of BBMRI-ERIC operations in Finland. The Finnish biobank data can be accessed through the Fingenious^®^services (https://site.fingenious.fi/en/) managed by FINBB.

This Estonian Biobank study was funded by European Union through the European Regional Development Fund Project No. 2014-2020.4.01.15-0012 GENTRANSMED. Data analysis was carried out in part in the High-Performance Computing Center of University of Tartu.

We acknowledge the Estonian Biobank research team: Andres Metspalu, Lili Milani, Reedik Mägi, Mari Nelis, and Georgi Hudjashov, giving them credit for data collection, genotyping, QC, and imputation.

## Supplementary Text

### Ethical approvals

The deCODE study was approved by the Icelandic National Bioethics Committee (VSN-15-169). The North West Research Ethics Committee reviewed and approved UK Biobank’s scientific protocol and operational procedures (REC reference no.: 06/MRE08/65).

Approval of the Copenhagen Hospital Biobank Reproductive Health Study (CHB-RHS) was obtained from the Danish National Committee on Health Research Ethics (NVK-1805807) and the Capital Region Data Protection Agency (P-2019-49).

All study participants provided a signed informed consent, and the study protocol has been approved by the administrative board of the Norwegian Mother, Father and Child Cohort Study, led by the Norwegian Institute of Public Health. The establishment of MoBa and initial data collection was based on a license from the Norwegian Data Protection Agency and approval from The Regional Committee for Medical Research Ethics. The study was approved by the Norwegian Regional Committee for Medical and Health Research Ethics South-East (2015/2425) and by the Swedish Ethical Review Authority (Dnr 2022-03248-01).

Participants in FinnGen provided informed consent for biobank research on basis of the Finnish Biobank Act. Alternatively, separate research cohorts, collected before the Finnish Biobank Act came into effect (in September 2013) and the start of FinnGen (August 2017) were collected on the basis of study-specific consent and later transferred to the Finnish biobanks after approval by Fimea, the National Supervisory Authority for Welfare and Health. Recruitment protocols followed the biobank protocols approved by Fimea. The Coordinating Ethics Committee of the Hospital District of Helsinki and Uusimaa (HUS) approved the FinnGen study protocol (number HUS/990/2017). The FinnGen study is approved by the Finnish Institute for Health and welfare (approval number THL/2031/6.02.00/2017, amendments THL/1101/5.05.00/2017, THL/341/6.02.00/2018, THL/2222/6.02.00/2018, THL/283/6.02.00/2019 and THL/1721/5.05.00/2019), the Digital and Population Data Service Agency (VRK43431/2017-3, VRK/6909/2018-3 and VRK/4415/2019-3), the Social Insurance Institution (KELA) (KELA 58/522/2017, KELA 131/522/2018, KELA 70/522/2019 and KELA 98/522/2019) and Statistics Finland (TK-53-1041-17).

The activities of the EstBB are regulated by the Human Genes Research Act, which was adopted in 2000 specifically for the operations of the EstBB. All Estonian Biobank participants have signed a broad informed consent form and analyses were carried out under ethical approval 1.1-12/624 from the Estonian Committee on Bioethics and Human Research (Estonian Ministry of Social Affairs) and data release N05 from the EstBB.

### Supplementary Figures

**Supplementary Figure 1.**
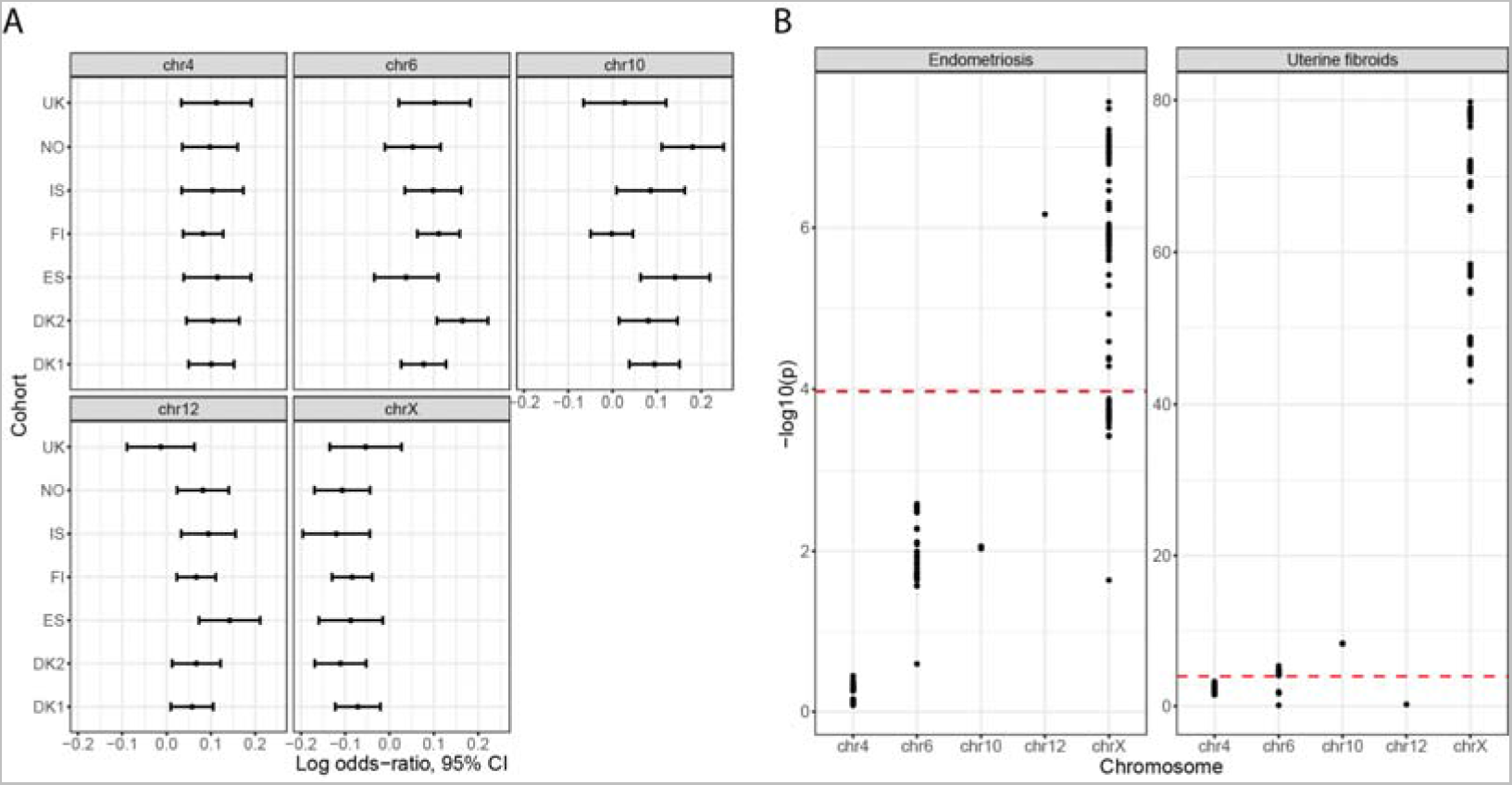
(A) Effect sizes in each cohort for the postpartum hemorrhage lead variants, which were largely similar. (B) Genome-wide significant variants from the postpartum hemorrhage analysis are also associated with endometriosis and uterine fibroids. The red line indicates the Bonferroni corrected p-value threshold (p < 0.05/(2 loci * 234 variants) = 0.0001).

**Supplementary Figure 2.**
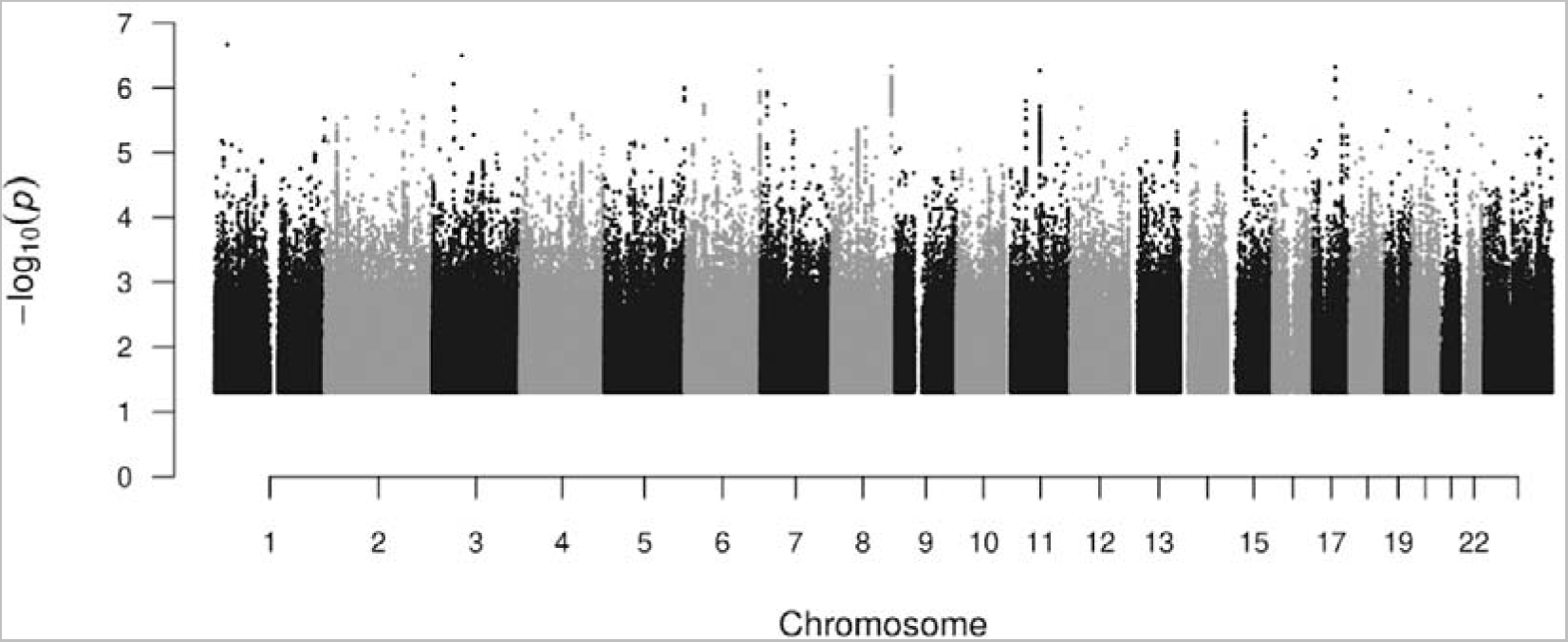
Manhattan plot for Early bleeding, all outcomes.

**Supplementary Figure 3.**
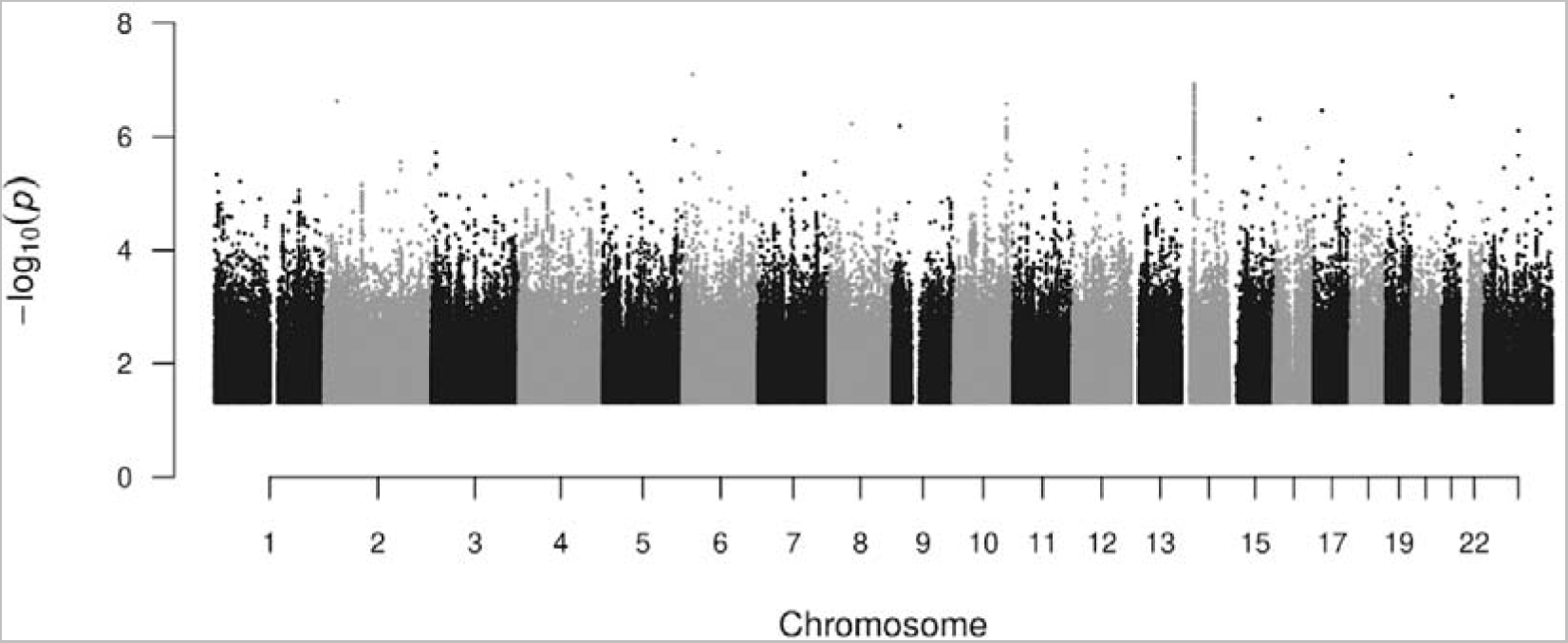
Supplementary Figure 2: Manhattan plot for Early bleeding, ending in live birth.

**Supplementary Figure 4.**
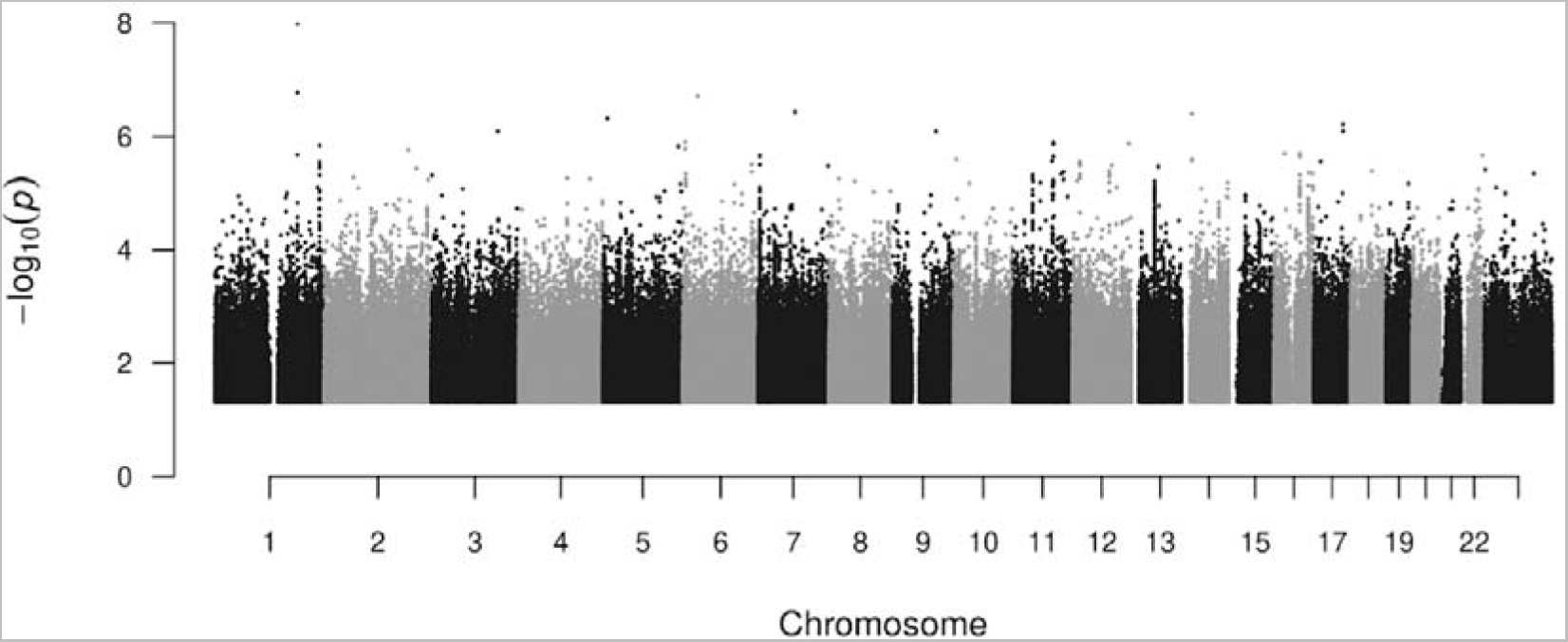
Manhattan plot for antepartum hemorrhage

**Supplementary Figure 5.**
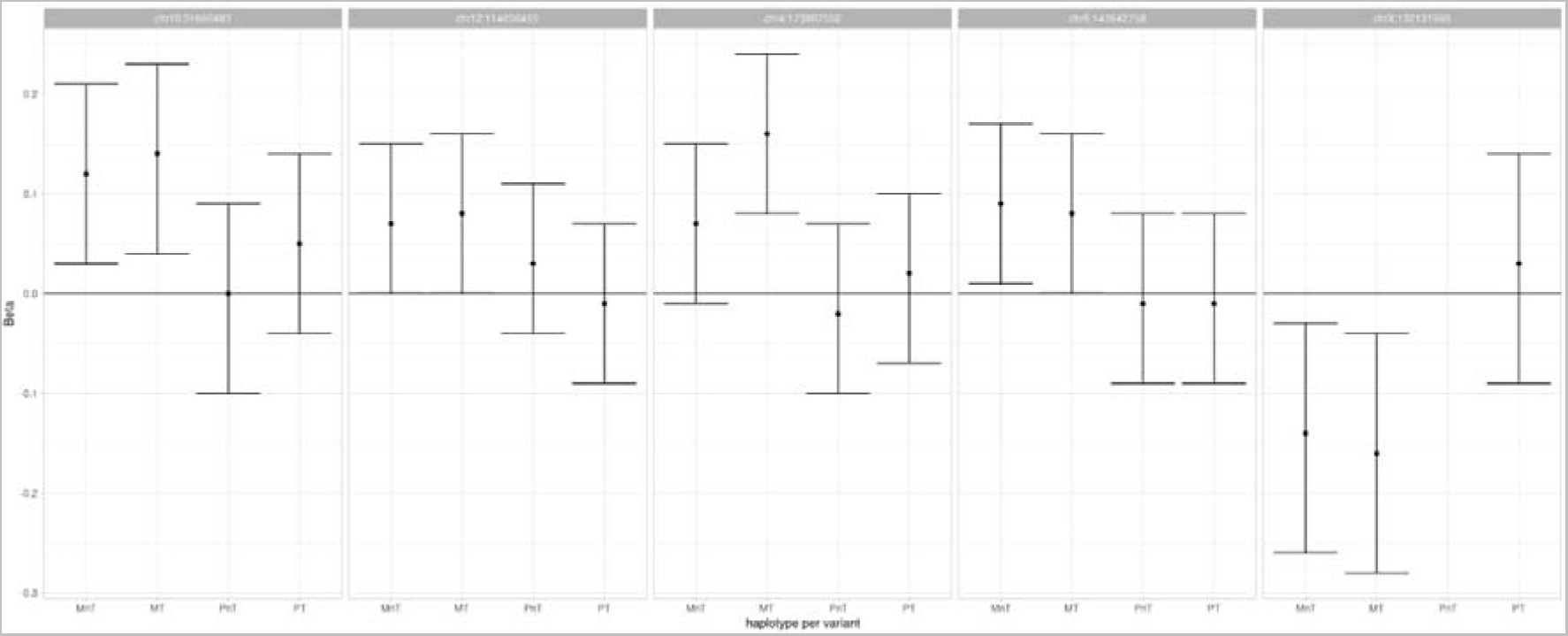
Haplotype analysis of the five PPH associated variants in the MoBa and deCODE cohorts. Results suggest that that effect is mediated through the maternal genome. Mnt: maternal non-transmitted; MT: maternal transmitted; PnT: paternal non-transmitted; PT: paternal transmitted

### Supplementary Tables

**Supplementary Table 1.**
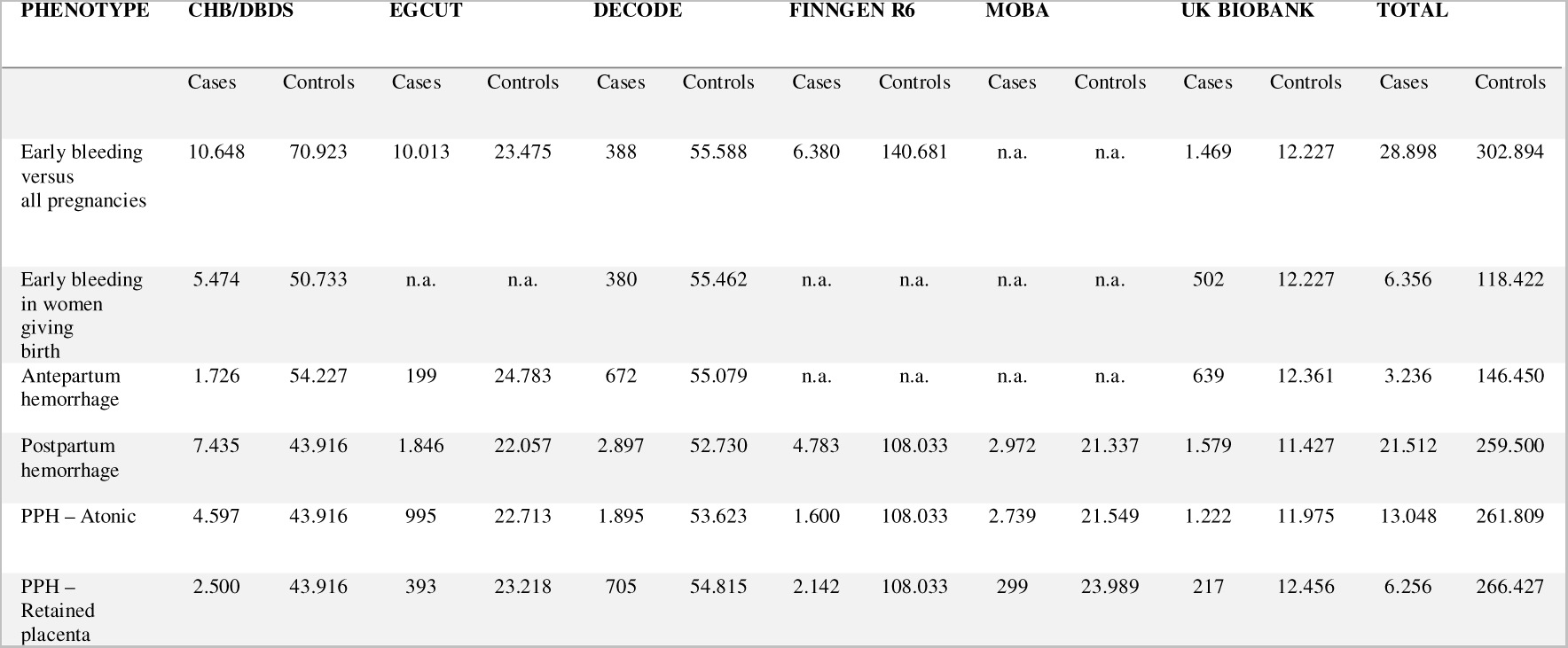
The contribution from each of the six Northern European cohorts. n.a. indicates the phenotype was not available in the cohort.

**Supplementary Table 2.** Effect sizes (95% CI and P-value) for the lead variants of PPH for the atoni and retained placenta subtypes, respectively.

| CHR | RSID | ATONI | RETAINED PLACENTA | CASE-CASE GWAS |
| --- | --- | --- | --- | --- |
| 4 | rs13141656 | 1.10 (1.08-1.13; 9.6e-16) | 1.12 (1.06-1.19; 0.00019) | 1.04 (0.99-1.09; 0.09) |
| 6 | rs12195857 | 1.10 (1.08-1.12; 2e-14) | 1.12 (1.06-1.17; 4.8e-05) | 1.03 (0.98-1.08; 0.26) |
| 10 | rs11591307 | 1.08 (1.05-1.10; 1.4e-09) | 1.10 (1.02-1.18; 0.019) | 1.05 (0.99-1.10; 0.08) |
| 12 | rs11067228 | 1.07 (1.05-1.10; 5e-09) | 1.08 (1.04-1.13; 0.00053) | 1.01 (0.97-1.06; 0.56) |
| X | rs2747025 | 0.91 (0.88-0.95; 3.1e-08) | 0.92 (0.90-0.95; 2.1e-10) | 1.05 (0.99-1.10; 0.05) |

**Supplementary Table 3.** LDSC statistics and inflation metrics.

| PHENOTYPE | LDSC H2,<br>OBSERVED SCALE<br>(SE) | LDSC<br>INTERCEP<br>T | RATIO | MEAN<br>CHI^2 |
| --- | --- | --- | --- | --- |
| Early bleeding in pregnancy, all outcomes | 0.0144 (0.0017) | 1.005 | 0.0688 | 1.08 |
| Early bleeding in pregnancy, live birth as outcome | 0.009 (0.0035) | 0.9929 | <0 | 1.02 |
| Antepartum hemorrhage | 0.0005 (0.0029) | 0.9975 | NA | 1 |
| Postpartum hemorrhage | 0.0149 (0.002) | 1.0175 | 0.176 | 1.10 |
| Postpartum hemorrhage due to atony | 0.0101 (0.0018) | 1.0092 | 0.1437 | 1.06 |
| Postpartum hemorrhage due to retained placenta | 0.0088 (0.0018) | 0.99 | <0 | 1.05 |

**Supplementary Table 4:** Variants previously associated with traits in the GWAS catalog (p <5·10^-8^), and variants in strong LD (r^2^ > 0.8) with other variants.

| CHROMOSOME | LEAD VARIANT | GWAS TRAIT, AS LISTED IN THE GWAS CATALOG | VARIANT IN LD | R<sup>2</sup> |
| --- | --- | --- | --- | --- |
| 6 | rs12195857 | Educational attainment | rs12200809 | 0.83 |
| 10 | rs11591307 | Uterine fibroids | rs11008551 | 0.98 |
| 10 | rs11591307 | Uterine fibroids | rs10508765 | 0.96 |
| 10 | rs11591307 | Uterine leiomyomata | rs7090544 | 0.97 |
| 12 | rs11067228 | Heel bone mineral density | rs11067228 | 1 |
| 12 | rs11067228 | Prostate-specific antigen levels | rs11067228 | 1 |
| 12 | rs11067228 | Serum prostate-specific antigen levels | rs11067228 | 1 |
| X | rs2747025 | Endometriosis | rs5933091 | 1 |
| X | rs2747025 | Serum creatinine levels | rs5933079 | 1 |
| X | rs2747025 | Uterine fibroids | rs5930554 | 1 |
| X | rs2747025 | Uterine fibroids | rs12392108 | 1 |
| X | rs2747025 | Uterine leiomyomata | rs5930554 | 1 |

**Supplementary Table 5.**
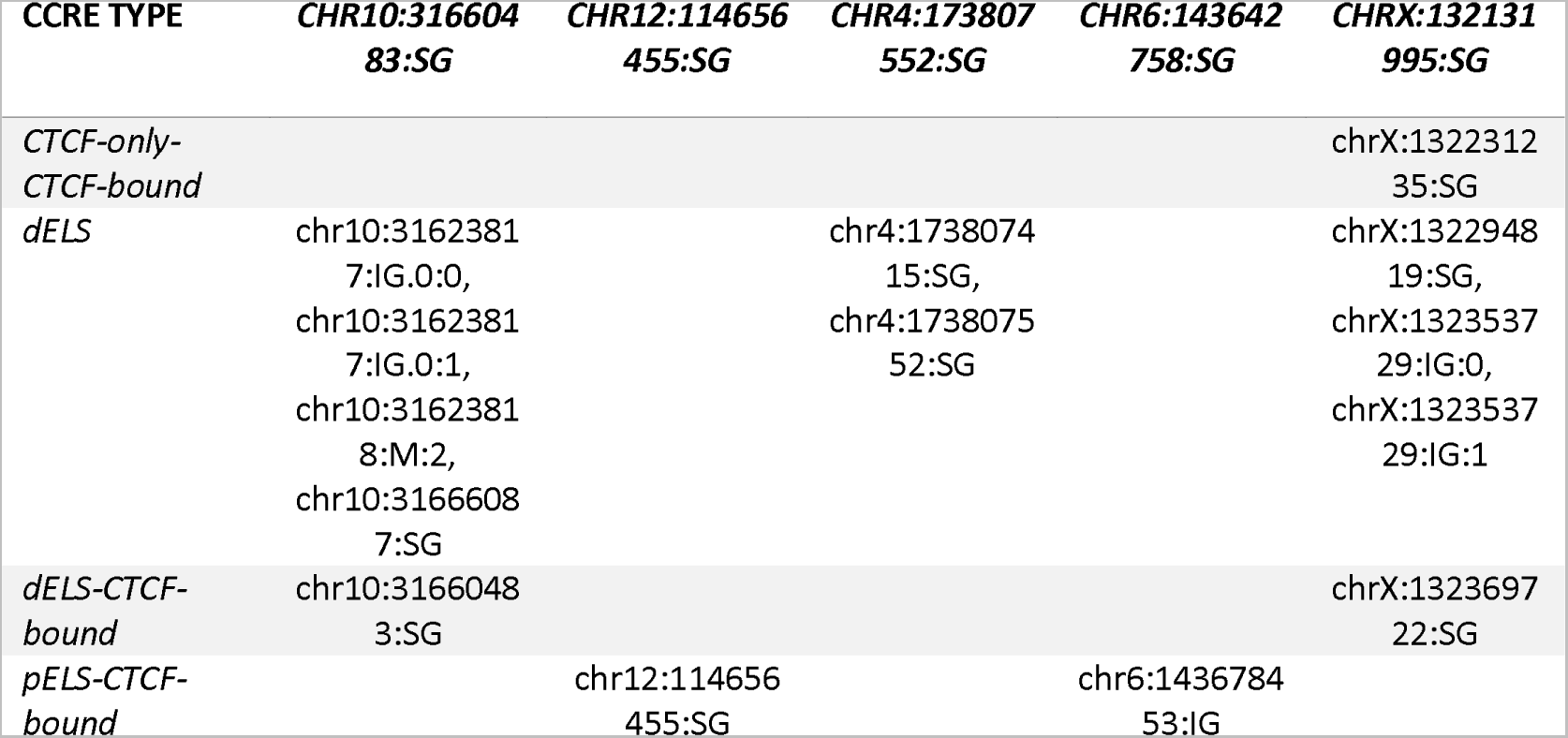
Post-partum hemorrhage (PPH) lead association sequence variants along with their correlated variants (r2>0.80) intersect with enhancer-like sequences (ELS) and CTCF binding sites as defined in ENCODÉs encyclopaedia of candidate cis-regulatory elements (cCRE).

**Supplementary Table 6.**
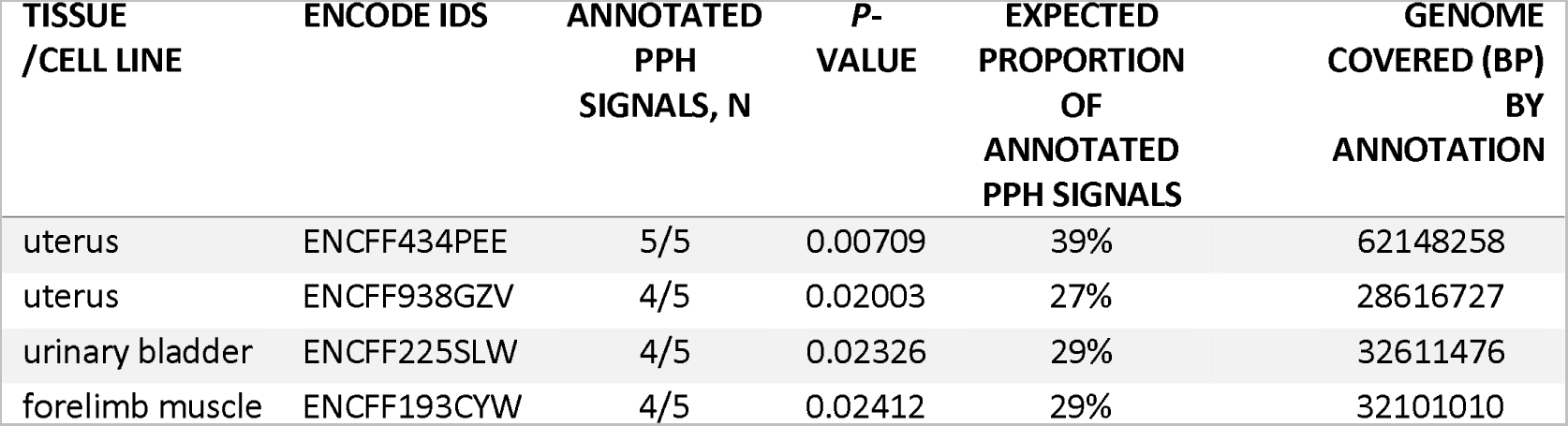
Enriched cCRE by Samples: PPH signals were tested for enrichment within cCREs of 1519 different samples representing 421 different tissues or cell types (UBERON and CL IDs). Shown are nominally significant results (P<0.05).

**Supplementary Table 7.** GWA signals in cCREs: PPH signals were nominally enriched within cCREs found in four (out of 1519) samples representing three different tissues (uterus, urinary bladder and forelimb muscle). Shown are the lead sequence variants (columns) or their correlated variants that were found in overlap with cCREs as defined in each of the five samples.

| TISSUE<br>/CELL<br>LINE | ENCODE IDS | CHR10:31660<br>483:SG | CHR12:1146<br>56455:SG | CHR4:1738<br>07552:SG | CHR6:1436<br>42758:SG | CHRX:1321<br>31995:SG |
| --- | --- | --- | --- | --- | --- | --- |
| uterus | **uterus**<br>**(ENCFF434PE**<br>**E)** | chr10:316238<br>17:IG.0:0,<br>chr10:316238<br>17:IG.0:1,<br>chr10:316238<br>18:M:2,<br>chr10:316604<br>83:SG | chr12:11465<br>6455:SG | chr4:17380<br>7415:SG,<br>chr4:17380<br>7552:SG | chr6:14367<br>8453:IG | chrX:13236<br>9722:SG |
| uterus | **uterus**<br>**(ENCFF938G**<br>**ZV)** | chr10:31623<br>817:IG.0:0,<br>chr10:31623<br>817:IG.0:1,<br>chr10:31623<br>818:M:2,<br>chr10:31660<br>483:SG | chr12:11465<br>6455:SG | chr4:17380<br>7415:SG,<br>chr4:17380<br>7552:SG | chr6:14367<br>8453:IG |  |
| urinary<br>bladder | **urinary**<br>**bladder**<br>**(ENCFF225SL**<br>**W)** | chr10:316238<br>17:IG.0:0,<br>chr10:316238<br>17:IG.0:1,<br>chr10:316238<br>18:M:2,<br>chr10:316604<br>83:SG | chr12:11465<br>6455:SG | chr4:17380<br>7415:SG,<br>chr4:17380<br>7552:SG | chr6:14367<br>8453:IG |  |
| forelimb<br>muscle | **forelimb**<br>**muscle**<br>**(ENCFF193CY**<br>**W)** | chr10:316238<br>17:IG.0:0,<br>chr10:316238<br>17:IG.0:1,<br>chr10:316238<br>18:M:2 |  | chr4:17380<br>7415:SG,<br>chr4:17380<br>7552:SG | chr6:14367<br>8453:IG | chrX:13236<br>9722:SG |

**Supplementary Table 8.** Enriched epimap: PPH signals were tested for enrichment within enhancers (A/G) as defined in 833 samples by Epimap. Shown are nominally significant results (P<0.05).

| TISSUE<br>/CELL LINE | EPIMAP<br>IDS | ANNOTATED<br>PPH<br>SIGNALS, N | P-VALUE | EXPECTED<br>PROPORTION<br>OF<br>ANNOTATED<br>PPH SIGNALS | GENOME<br>COVERED (BP)<br>BY<br>ANNOTATION |
| --- | --- | --- | --- | --- | --- |
| UTERUS | BSS01884 | 4/5 | 0.0085 | 22% | 50940614 |
| MESENDODERM DERIV | BSS01263 | 3/5 | 0.013 | 12% | 22150855 |

**Supplementary Table 9.** Predicted gene targets, Epimap: PPH signals were most strongly enriched among enhancers (Active/Genic) found in uterus and three other tissues in Epimap*. Shown is the intersection for each PPH signal (lead variant and their correlated variants given r2>0.80) with enhancers in each of the most strongly enriched tissues where P<0.05, nominally significant, and the predicted gene target for those enhancers.

| TISSUE /<br>CELL-TYPE | **10P11.22**<br>**(CHR10:31660**<br>**483:SG)** | **12Q24.21**<br>**(CHR12:114656**<br>**455:SG)** | **6Q24.2**<br>**(CHR6:14364**<br>**2758:SG)** | **XQ26.2**<br>**(CHRX:132131995:**<br>**SG)** | **4Q34.1**<br>**(CHR4:17380**<br>**7552:SG)** |
| --- | --- | --- | --- | --- | --- |
| UTERUS |  | TBX3<br>(chr12:114656<br>455:SG) |  | FRMD7<br>(chrX:132120786:S<br>G,chrX:132126844:<br>SG), RAP2C<br>(chrX:132266705:S<br>G) |  |
| MESENDO<br>DERM_DE<br>RIV |  | TBX3<br>(chr12:114656<br>455:SG) |  | FRMD7<br>(chrX:132120786:S<br>G) |  |

**Supplementary Table 10.** Haplotype effect estimates for the five PPH associated loci.

|  |  | MOBA |  |  | DECODE |  |  | META |  |  |  |  |
| --- | --- | --- | --- | --- | --- | --- | --- | --- | --- | --- | --- | --- |
|  | Haplotype Variant | effect | 95%CI | P | effect | 95%CI | P | effect | 95%CI | P | P_het | I2 |
| MT | chr10:31660483 | 0.13 | (0.03;0.22) | 0.0099 | 0.23 | (-0.08;0.54) | 0.15 | 0.14 | (0.04;0.23) | 0.0039 | 0.53 | 0 |
|  | 12:114656455 | 0.07 | (-0.01;0.15) | 0.095 | 0.2 | (-0.05;0.46) | 0.11 | 0.08 | (0.00;0.16) | 0.038 | 0.32 | 0.9 |
|  | 4:173807552 | 0.16 | (0.08;0.25) | 0.00019 | 0.13 | (-0.16;0.42) | 0.37 | 0.16 | (0.08;0.24) | 0.00012 | 0.83 | 0 |
|  | 6:143642758 | 0.06 | (-0.03;0.15) | 0.17 | 0.24 | (-0.02;0.50) | 0.071 | 0.08 | (-0.00;0.16) | 0.060 | 0.20 | 38.9 |
|  | X:132131995 | -0.16 | (-0.28;-0.04) | 0.0099 | -0.13 | (-0.56;0.31) | 0.57 | -0.16 | (-0.28;-0.04) | 0.0084 | 0.88 | 0 |
| MnT | 10:31660483 | 0.12 | (0.03;0.22) | 0.012 | 0.09 | (-0.26;0.44) | 0.60 | 0.12 | (0.03;0.21) | 0.010 | 0.88 | 0 |
|  | 12:114656455 | 0.06 | (-0.02;0.14) | 0.17 | 0.25 | (-0.02;0.52) | 0.066 | 0.07 | (-0.00;0.15) | 0.065 | 0.17 | 45.9 |
|  | 4:173807552 | 0.06 | (-0.02;0.15) | 0.15 | 0.12 | (-0.18;0.43) | 0.43 | 0.07 | (-0.01;0.15) | 0.11 | 0.72 | 0 |
|  | 6:143642758 | 0.07 | (-0.01;0.16) | 0.095 | 0.28 | (0.00;0.55) | 0.046 | 0.09 | (0.01;0.17) | 0.028 | 0.16 | 49 |
|  | X:132131995 | -0.17 | (-0.30;-0.05) | 0.0048 | 0.06 | (-0.25;0.36) | 0.71 | -0.14 | (-0.26;-0.03) | 0.013 | 0.17 | 48 |
| PT | 10:31660483 | 0.04 | (-0.06;0.13) | 0.47 | 0.18 | (-0.13;0.50) | 0.25 | 0.05 | (-0.04;0.14) | 0.3 | 0.38 | 0 |
|  | 12:114656455 | -0.03 | (-0.11;0.05) | 0.48 | 0.17 | (-0.08;0.43) | 0.18 | -0.01 | (-0.09;0.07) | 0.80 | 0.13 | 55.4 |
|  | 4:173807552 | 0.02 | (-0.06;0.11) | 0.62 | -0.07 | (-0.36;0.23) | 0.66 | 0.02 | (-0.07;0.10) | 0.72 | 0.58 | 0 |
|  | 6:143642758 | -0.04 | (-0.13;0.05) | 0.38 | 0.28 | (0.02;0.54) | 0.035 | -0.01 | (-0.09;0.08) | 0.88 | 0.023 | 80.8 |
|  | X:132131995 | 0.02 | (-0.09;0.14) | 0.69 | 0.04 | (-0.38;0.46) | 0.85 | 0.03 | (-0.09;0.14) | 0.66 | 0.94 | 0 |
| PnT | 10:31660483 | -0.01 | (-0.11;0.09) | 0.80 | 0.11 | (-0.25;0.47) | 0.55 | 0 | (-0.10;0.09) | 0.93 | 0.52 | 0 |
|  | 12:114656455 | 0.04 | (-0.04;0.12) | 0.37 | -0.02 | (-0.32;0.27) | 0.89 | 0.03 | (-0.04;0.11) | 0.41 | 0.71 | 0 |
|  | 4:173807552 | -0.01 | (-0.10;0.08) | 0.83 | -0.13 | (-0.48;0.21) | 0.44 | -0.02 | (-0.10;0.07) | 0.69 | 0.49 | 0 |
|  | 6:143642758 | 0 | (-0.09;0.08) | 0.94 | -0.04 | (-0.34;0.27) | 0.82 | -0.01 | (-0.09;0.08) | 0.90 | 0.84 | 0 |
|  | 12:114656455 | 0.04 | (-0.04;0.12) | 0.37 | -0.02 | (-0.32;0.27) | 0.89 | 0.03 | (-0.04;0.11) | 0.41 | 0.71 | 0 |
|  | 4:173807552 | -0.01 | (-0.10;0.08) | 0.83 | -0.13 | (-0.48;0.21) | 0.44 | -0.02 | (-0.10;0.07) | 0.69 | 0.49 | 0 |
|  | 6:143642758 | 0 | (-0.09;0.08) | 0.94 | -0.04 | (-0.34;0.27) | 0.82 | -0.01 | (-0.09;0.08) | 0.90 | 0.84 | 0 |

**Supplementary Table 11.** Data sets used to generate summary statistics for genetic correlation analysis. ICE: deCODE genetics; GBR: UK Biobank; DNK: CHB/DBDS; FIN: FinnGen r7.

| *Trait* | *PPH* | *EB* | *Analysis* | *cases* | *ctrls* | *Datasets/Reference* |
| --- | --- | --- | --- | --- | --- | --- |
| *Pregnancy loss* |  | *x* | *meta* | *49.996* | *174.109* | *PMID: 33239672* |
| *Uterine fibroids* | *x* | *x* | *meta* | *49.418* | *893.310* | *ICE_GBR_FIN* |
| *Endometriosis* | *x* | *x* | *meta* | *25.137* | *881.619* | *ICE_GBR_DNK_FIN* |
| *Gestational diabetes* | *x* |  | *meta* | *9.507* | *928.959* | *ICE_GBR_FIN* |
| *Birth weight, fetal* | *x* |  | *meta* | *419.140* |  | *PMID: 34282336* |
| *Birth weight, maternal* | *x* |  | *meta* | *268.129* |  | *PMID: 34282336* |
| *Gestational age, maternal* | *x* |  | *GWAS* | *59.496* |  | *ICE* |
| *Gestational age, fetal* | *x* |  | *GWAS* | *125.228* |  | *ICE* |
| *Body mass index* | *x* | *x* | *meta* | *509.458* |  | *ICE_GBR* |
| *Smoking* |  |  | *meta* | *313.810* | *581.902* | *ICE_GBR_FIN* |
| *Alcohol dependence* |  |  | *meta* | *29.557* | *707.411* | *ICE_GBR* |
| *Education years* |  |  | *GWAS* | *403.567* |  | *GBR* |
| *Risk taking* |  |  | *GWAS* | *107.158* | *323.889* | *GBR* |
| *HDL Cholesterol* |  |  | *meta* | *548.375* |  | *ICE_GBR* |
| *Type 2 diabetes* | *x* | *x* | *meta* | *88.062* | *916.726* | *ICE_GBR_FIN* |
| *Systolic blood pressure* | *x* | *x* | *meta* | *508.767* |  | *ICE_GBR* |
| *Diastolic blood pressure* | *x* | *x* | *meta* | *508.764* |  | *ICE_GBR* |
| *Hypertension* | *x* | *x* | *meta* | *347.202* | *901.986* | *ICE_GBR_DNK_FIN* |
| *Coronary artery disease* | *x* | *x* | *meta* | *172.831* | *1.051.258* | *ICE_GBR_DNK_FIN* |
| *Myocardial infarction* | *x* | *x* | *meta* | *76.141* | *1.121.254* | *ICE_GBR_DNK_FIN* |
| *Atrial fibrillation* | *x* | *x* | *meta* | *108.925* | *970.342* | *ICE_GBR_DNK_FIN* |
| *Heart failure* | *x* | *x* | *meta* | *88.888* | *1.184.381* | *ICE_GBR_DNK_FIN* |
| *Stroke ischemic* |  | *x* | *meta* | *83.860* | *1.049.069* | *ICE_GBR_FIN PMID: 29531354* |
| *Stroke intracerebral hemorrhage* |  | *x* | *daisy* | *12.224* | *339.944* | *ICE* |
| *Post traumatic stress disorder* | *x* | *x* | *meta* | *5.067* | *1.023.714* | *ICE_GBR_USA_FIN* |
| *Stress* | *x* | *x* | *GWAS* | *12.655* | *19.225* | *PMID: 31116379* |
| *Major depression* | *x* | *x* | *meta* | *59.851* | *113.154* | *PMID: 29700475* |
| *Asthma* | *x* | *x* | *meta* | *125.769* | *842.132* | *ICE_GBR_FIN* |
| *Hypothyroidism* | *x* | *x* | *meta* | *62.503* | *787.353* | *ICE_GBR_FIN* |
| *Cholelithiasis* | *x* | *x* | *meta* | *23.087* | *721.958* | *ICE_GBR* |

**Supplementary Table 12:** Effect sizes for the standardized polygenic risk scores of PPH and birth weight.

| **VARIABLE** | **EFFECT SIZE, LOG-ODDS** | **95% CI** |
| --- | --- | --- |
| PPH | 1.08 | 1.05;1.11 |
| Birth weight | 1.14 | 1.10; 1.17 |

**Supplementary Table 13:** Cohort definitions.

| PHENOTYPE | CODES | EXCLUSION | NOTE |
| --- | --- | --- | --- |
| Bleeding in early pregnancy ending in any outcome | ICD8:<br>ICD9:<br>ICD10:O20.* | Known coagulation disorders (D66-D69, O46.0, O67.0) |  |
| Bleeding in early pregnancy leading to live birth | ICD8:<br>ICD9:<br>ICD10:O20.* | Known coagulation disorders (D66-D69, O46.0, O67.0) | Code must have occurred within the gestational period leading to a stillbirth or livebirth. |
| Antepartum hemorrhage | ICD8:<br>ICD9:<br>ICD10: O46 | Known coagulation disorders (D66-D69, O46.0, O67.0) |  |
| Postpartum hemorrhage | ICD8:<br>ICD9:<br>ICD10: O72<br>Post-2012 (DK only):<br>ICD10: O72 (with >500mL blood loss) | Known coagulation disorders (D66-D69, O46.0, O67.0)<br>Multifold pregnancy Cesarean sectio | In Denmark, it has since 2012 been mandatory to report the amount of blood lost during birth along with the O72 diagnosis. |
| PPH due to atony | ICD8:<br>ICD9:<br>ICD10: O72.1, and no trauma/retained tissue<br>Post-2012 (DK only):<br>ICD10: O72 (with >500mL blood loss), and no trauma/retained tissue | Known coagulation disorders (D66-D69, O46.0, O67.0)<br>Multifold pregnancy Cesarean sectio |  |
| PPH due to retained placenta | ICD8:<br>ICD9:<br>ICD10: O72.2<br>Post-2012 (DK only):<br>ICD10: O72 (with >500mL blood loss) + O73 | Known coagulation disorders (D66-D69, O46.0, O67.0)<br>Multifold pregnancy Cesarean sectio |  |

